# Ischemic Stroke Detection, Segmentation, and Volume Estimation using Multi-sequence MRI Data with Missing Sequences

**DOI:** 10.64898/2026.09.12.26362795

**Authors:** Zhicheng Lu, Shahadat Uddin, Sergio Uribe, Sam White, Rodrigo Tomazini Martins, Shayne Chau, Abu Syed Md. Mosaddek, Md. Siddiqul Islam, Nusratun Nahar, AKM Azad, K. M. Nazmul Hossain, Habib Sadat Choudhury, K.M. Rakibul Hasan, Saad Aloteibi, Nabil Mosaddek, Samia Rahman, Md. Mostaque Hossain, K.M. Mehedi Hasan Sizar, Claudio Angione, Pietro Liò, Md Tauhidul Islam, Mohammad Ali Moni

## Abstract

Stroke remains one of the leading causes of disability and mortality worldwide, where timely and accurate diagnosis is critical for guiding treatment and improving patient outcomes. However, a global shortage of trained clinicians and radiologists continues to limit rapid and reliable interpretation of neuroimaging, particularly in resource-constrained settings. Artificial intelligence (AI) has emerged as a promising solution to this challenge by enabling efficient analysis of medical images. Here we present an Integrated Stroke Diagnosis System for MRI (ISDS-MRI), a unified framework that leverages graph neural networks and sequence-specific feature modeling for comprehensive ischemic stroke analysis. This framework is designed to detect ischemic stroke, segment lesions, and estimate lesion volume from multi-sequence MRI data, while accommodating incomplete combinations of MRI sequences. To ensure generalizability, we evaluate our approach across multiple publicly available MRI datasets and introduce a newly curated dataset, BGD-MRIS, comprising 532 MRI scans from three hospitals in Bangladesh. This newly curated dataset provides a multi-center MRI cohort from a resource-constrained setting, offering an additional test bed for evaluating stroke AI across heterogeneous clinical imaging protocols. Experimental results demonstrate that ISDS-MRI achieves a Dice score of 0.725 for lesion segmentation, a AUC of 0.962, and a lesion volume estimation relative error of 8.4%, outperforming comparison methods by 3.2% in Dice score and 2.6% in detection performance, while reducing volume estimation relative error by 1.9%. These results highlight the robustness and clinical potential of ISDS-MRI for scalable and comprehensive stroke diagnosis from MRI. The BGD-MRIS dataset will be publicly available at https://github.com/Zhicheng-Lu/stroke_mri.

## Introduction

Stroke is a leading cause of long-term disability and the second leading cause of mortality worldwide^1,2^. Ischemic stroke accounts for approximately 80–85% of all stroke cases^3^, affecting both developed and developing nations. In the United States, stroke ranks as the fifth leading cause of death^4^, resulting in roughly 140,000 fatalities annually and nearly one million individuals affected^5^. The burden is even greater in developing countries, where around 75% of stroke-related deaths and 85% of stroke-related disabilities worldwide occur^6^. For example, in Bangladesh, stroke is the second leading cause of mortality and the third leading cause of disability, representing one of the highest cerebrovascular disease burdens in South Asia^7^. Incidence disproportionately affects older populations, with rates among individuals aged over 75 reaching 1,200 per 100,000, compared to 150 per 100,000 in the general population^1^.

Early and accurate diagnosis of acute ischemic stroke, followed by timely intervention, can substantially improve clinical outcomes^8,9^. Clinical practice typically relies on computed tomography (CT) and magnetic resonance imaging (MRI) for diagnosis. While CT offers rapid acquisition and widespread availability, MRI provides superior soft-tissue contrast and higher sensitivity for detecting ischemic lesions^10,11^. In particular, diffusion-weighted imaging (DWI) MRI, which detects changes in the Brownian motion of water molecules in affected tissue, is highly sensitive and can identify ischemic stroke within minutes of onset in 50–95% of cases^12^. Despite these advantages, there is a critical global shortage of radiologists trained in MRI interpretation^13^.

To address these limitations, computer science researchers have partnered with clinicians to develop AI-based medical image analysis systems, including approaches for image enhancement^14^, lesion detection^15^, and lesion segmentation^16^. In stroke diagnosis, deep learning methods have demonstrated substantial potential for improving both accuracy and efficiency. MRI-based machine learning and deep learning techniques have achieved diagnostic performance comparable to that of expert clinicians^17–20^. Beyond classification, lesion segmentation and volume estimation provide clinically meaningful information that can guide patient management and inform prognosis. U-Net, introduced by Ronneberger *et al*.^21^, has become a foundational architecture for medical image segmentation, with numerous extensions and variants^22–29^ achieving performance on par with expert annotations. Nevertheless, three-dimensional multi-sequence MRI features remain under-explored, particularly for jointly supporting lesion detection, segmentation, and quantitative lesion volume estimation. Effective MRI-based stroke analysis can benefit from complementary information across multiple sequences. However, the availability of individual sequences varies substantially across clinical examinations and public datasets because of differences in acquisition protocols, patient tolerance, acquisition time, and institutional practice. Existing stroke MRI datasets therefore contain highly heterogeneous and often non-overlapping sequence combinations. For example, ISLES22^30^ provides ADC and DWI, whereas ATLAS R2.0^27^ provides T1w images. A framework that requires a fixed and complete set of sequences would consequently exclude a substantial proportion of available data. This motivates a sequence-flexible design that can operate using the subset of MRI sequences available for a given case. Although sequence synthesis techniques have been proposed^31–33^, their effectiveness in stroke remains limited.

In this work, we present ISDS-MRI, a simple yet effective framework for multi-sequence MRI stroke analysis (Figure 1). Each MRI sequence (ADC, DWI, FLAIR, T1w, or T2w) is processed by a dedicated sequence-specific sub-model with independently learned parameters (Supplementary Figure S1). For each case, only sub-models corresponding to the available sequences are activated, and their predictions are aggregated to generate the final output. Consequently, ISDS-MRI does not require every patient to have the same fixed set of MRI sequences and can directly accommodate incomplete sequence combinations without synthesizing missing images. Spatial features are extracted using 2D U-Net^21^, while three-dimensional contextual relationships are modeled using graph neural networks (GNNs) that connect semantically similar slices^34^. We further leverage segmentation-derived features for ischemic stroke detection, while lesion volume is calculated directly from the predicted segmentation masks and physical voxel dimensions.

**Figure 1.**
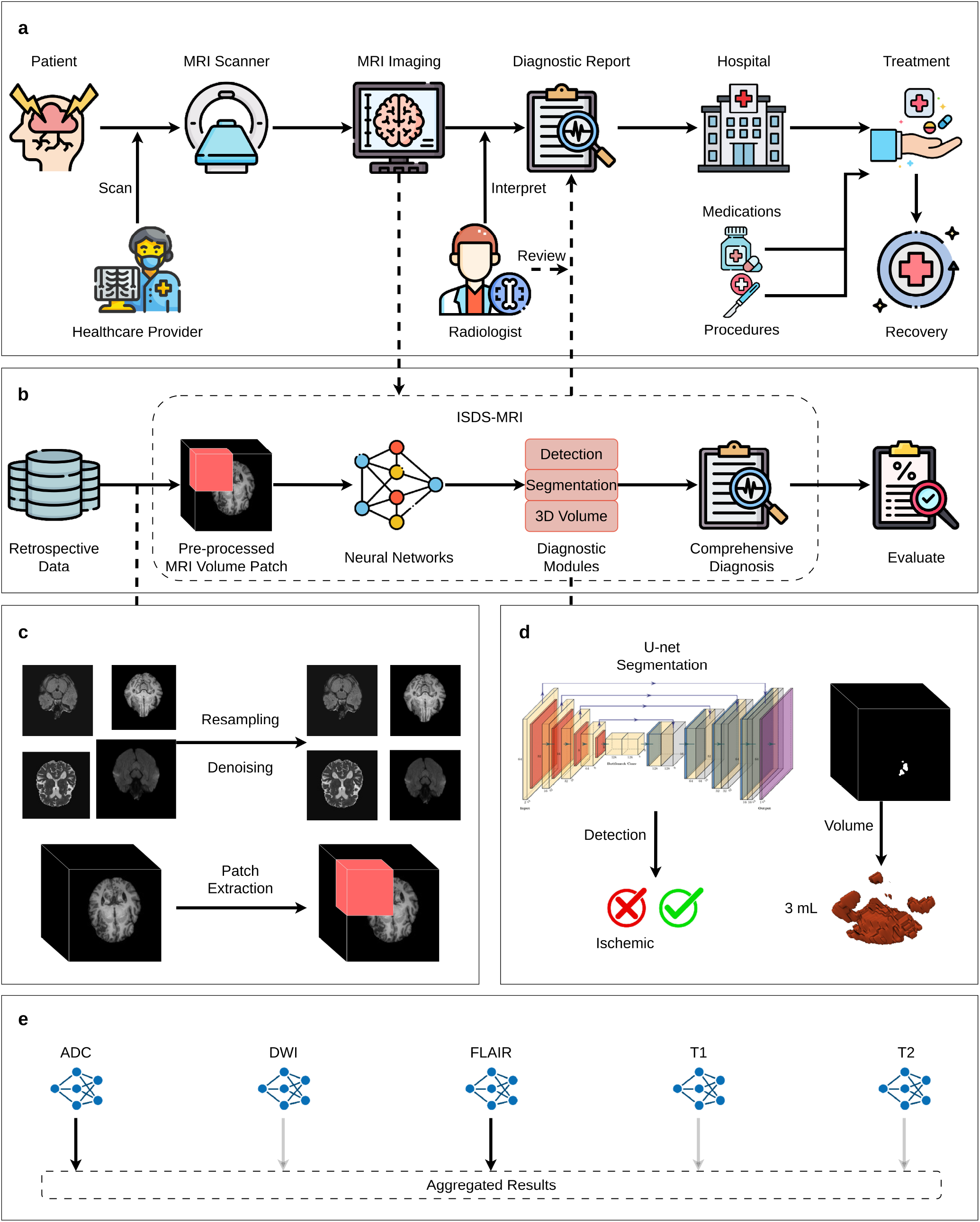
Overview of the proposed workflow. (a) Traditional clinical pathway: Suspected stroke patients undergo MRI scan, after which radiologists manually review the scans and generate diagnostic reports to guide subsequent treatment. (b) AI-enabled workflow: MRI images are automatically processed by the pre-trained ISDS-MRI, which provides a comprehensive diagnostic output including ischemic stroke detection, lesion segmentation, and 3D volume estimation, radiologists review and confirm the output from ISDS-MRI. (c) Pre-processing pipeline including resampling, denoising, and patch-volume reconstruction. (d) Multi-task diagnostics, including stroke detection, segmentation, and volume estimation. Model architecture design is described in Supplementary Figure S1 and Supplementary Table S2. Details of the deep learning model design, including StrokeGNN and transfer learning, are described in Section Methods.

To validate our framework, we introduce the Bangladesh Ischemic Stroke Dataset (BGD-MRIS), consisting of 532 MRI series from 115 patients collected across three medical centers in Bangladesh. To support reproducibility and future research, ISDS-MRI will be publicly released at https://github.com/Zhicheng-Lu/stroke_mri. Trained on multiple public datasets and evaluated on both public and private data, ISDS-MRI demonstrates competitive performance in detection and volume estimation while achieving state-of-the-art segmentation results. These findings underscore the potential of ISDS-MRI for clinical deployment in diverse healthcare environments.

## Results

### ISDS-MRI-enabled ischemic stroke lesion segmentation

To develop a generalized MRI-based stroke analysis framework, lesion segmentation is treated as a core component of ISDS-MRI, and the model is trained using multiple publicly available MRI datasets. SISS^35^ consists of 64 sub-acute ischemic stroke cases from the University Medical Center Schleswig-Holstein in Lübeck, Germany. Available MRI sequences include T1w, T2w, DWI, and FLAIR.. Ground-truth masks were created by two experienced experts based on the FLAIR sequence with additional information from the other 3 MRI sequences. SPES^35^ contains 50 cases acquired at the University Hospital of Bern between 2005 and 2013. Available sequences include T1w, T1c, T2w, DWI, and perfusion-derived sequences including cerebral blood flow (CBF), cerebral blood volume (CBV), time-to-peak (TTP), and time-to-maximum (Tmax). In this study, we retain T1w, T2w, and DWI to maintain consistency with the sequence set considered across datasets, while the perfusion-derived sequences are excluded from the present analysis. Lesions were labelled manually based on the DWI sequence. SISS and SPES together form the MICCAI ISLES2015 challenges. ISLES22^30^ dataset is part of ISLES 2022 challenge, the dataset has 400 cases with DWI and ADC sequences. ATLAS R2.0^27^ is an independent large-scale stroke lesion dataset containing T1w MRI and manually delineated lesion masks. The slice thickness distribution is illustrated in Supplementary Figure S2b.

To reduce inter-site variability across multi-centre MRI data, all scans are spatially standardized by resampling to a uniform in-plane resolution of 0.5× 0.5 mm, while preserving the original through-plane (slice) spacing. This approach harmonizes spatial resolution without introducing unnecessary interpolation artifacts in thick-slice acquisitions, thereby retaining acquisition-specific characteristics commonly encountered in real-world clinical practice. Consequently, datasets with finer through-plane resolution may contain more slices, providing ISDS-MRI with richer volumetric context. For computational efficiency, each MRI volume ℝ^*D*×*W* ×*H*^ is decomposed into overlapping sub-volumes of size ℝ ^30×256×256^. A minimum in-plane overlap of 32 pixels is maintained between neighboring patches to preserve spatial continuity. This patch-based training strategy reduces GPU memory requirements while enabling effective learning of three-dimensional representations of ischemic stroke lesions. Volumes containing fewer than 30 slices are kept intact without further partitioning.

Firstly, we test the performance of the proposed lesion segmentation model in a 5-fold cross-validation format. We augment datasets by following rules: SISS by a scaling factor of 30, SPES by a scaling factor of 20, and ISLES22 by a scaling factor of 3. The distribution of training, validation, and testing sets is described in Figure 2a (see Supplementary Figure S2a for details). All folds are constructed at the patient level before augmentation to ensure that augmented versions of the same case cannot appear across training, validation, and test sets. Data augmentation is applied only after the training/validation split, while the held-out test fold remains unaugmented. We alternatively use augmented MRI data from 4 out of 5 folds (i.e., 557×4 = 2228 cases) as training and validation sets with a ratio of 2000 training samples and 228 validation samples, and use the remaining fold with non-augmented 192 samples as the test set. Based on true positive (TP), true negative (TN), false positive (FP), and false negative (FN), we evaluate segmentation performance using the Dice score, Intersection over Union (IoU), precision, recall, Average Symmetric Surface Distance (ASSD), and Hausdorff Distance (HD).

**Figure 2.**
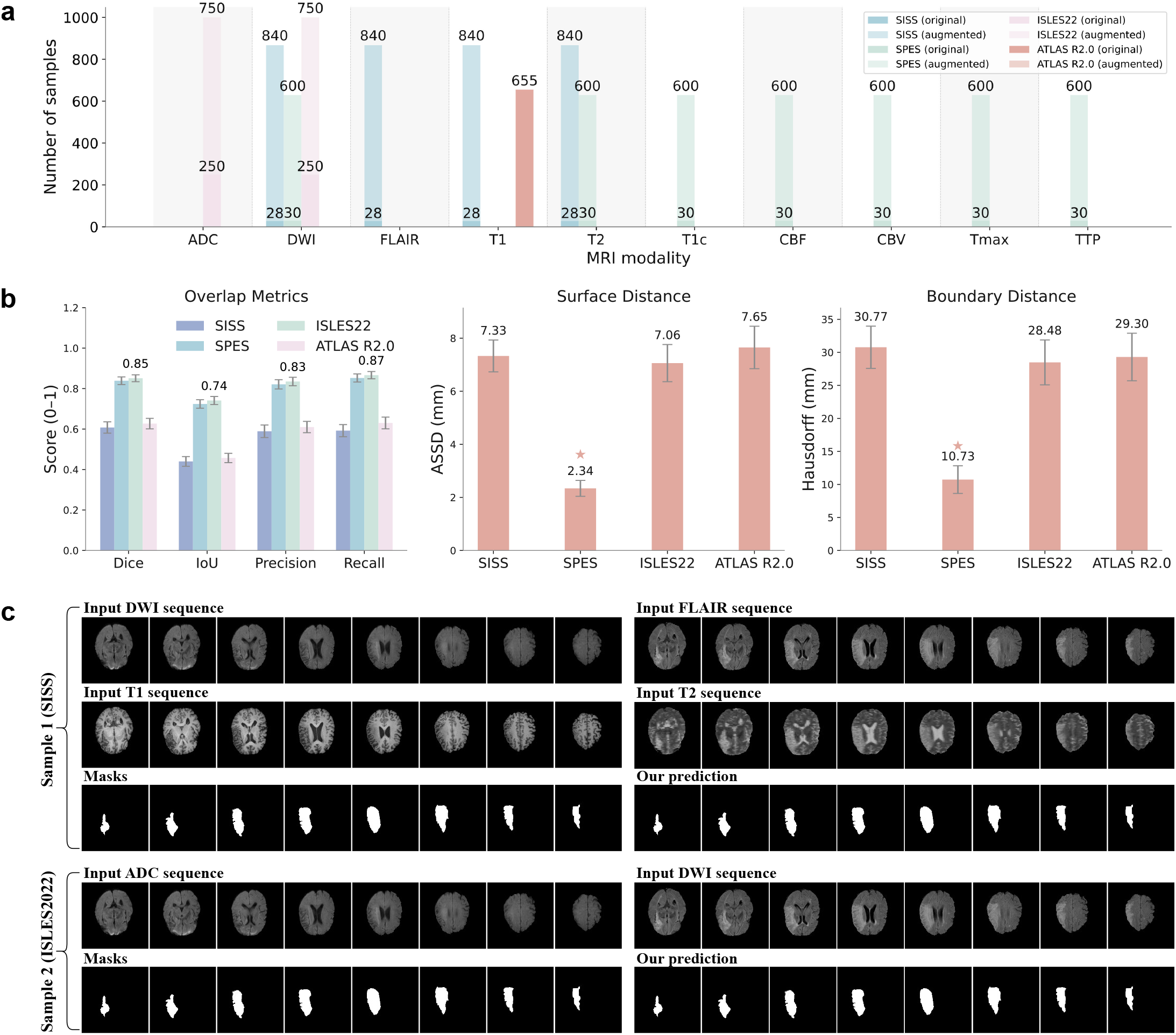
Experimental results of stroke lesion segmentation using cross-validation with fold 1 as the test set. (a) Dataset distribution for lesion segmentation training - SISS, SPES, ISLES22, and ATLAS R2.0 on 10 different MRI sequences. Note that numbers in the brackets are the dataset size after augmentation. (b) Average experimental results on average of 5-fold cross validation. (c) Sample experimental results from SISS and ISLES2022. Each sample includes MRI sequences, ground-truth masks, and our predicted segmentation map.

We then evaluate the proposed model using a 5-fold cross-validation. Overall, the model takes on average approximately 69 epochs (approximately 600 minutes) to converge based on the early stop criteria. Figure S2c displays the training and validation loss values when we use fold 4 as the test set in a 5-fold cross-validation approach. Training terminates at the 67^th^ epoch after the validation loss fails to improve for five consecutive epochs. Experimental results for the segmentation task in the cross-validation approach are shown in Figure 2b (see Supplementary Figure S2d for details). Our proposed model achieves an average Dice score of 72.50, with both SPES and ISLES22 attaining Dice scores above 0.80. Figure 2c displays subjective experimental results on 2 testing samples from 2 different datasets, including multimodal MRI sequences, labelled masks, and our prediction. From Figure 2c, qualitative inspection shows that the predicted lesion masks closely correspond to the reference annotations across representative cases from different datasets.

To further evaluate and validate the effectiveness of our proposed system, we compare it against state-of-the-art methods using the SISS and ATLAS R2.0 datasets. In this experiment, we reuse the four datasets mentioned above and apply data augmentation to SISS, SPES, and ISLES22 to increase the size and diversity of the training set. The dataset distribution used for comparison with existing methods is shown in Figure S3a. We randomly select one-eighth 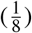 of the training data as the validation set. We use the same setup as in the cross-validation experiment. The training and validation losses across epochs are shown in Figure S3b. The training process completes in 74 epochs (approximately 700 minutes), with validation accuracy remaining stable throughout. We contrast our proposed method with several existing methods on SISS as follows: Kamnitsas *et al*.^22^, Feng *et al*.^36^, and Halme *et al*.^37^. Figure 3a (see Supplementary Figure S3c for details) compares our proposed method with state-of-the-art approaches on the SISS dataset in terms of Dice score, recall, Average Symmetric Surface Distance (ASSD), and Hausdorff Distance (HD). Our method achieves the highest Dice score of 0.6182, outperforming Kamnitsas *et al*.^22^ (0.59), Feng *et al*.^36^ (0.55), and Halme *et al*.^37^ (0.47). It also demonstrates the best performance in terms of ASSD and ranks among the top methods for recall and HD.

**Figure 3.**
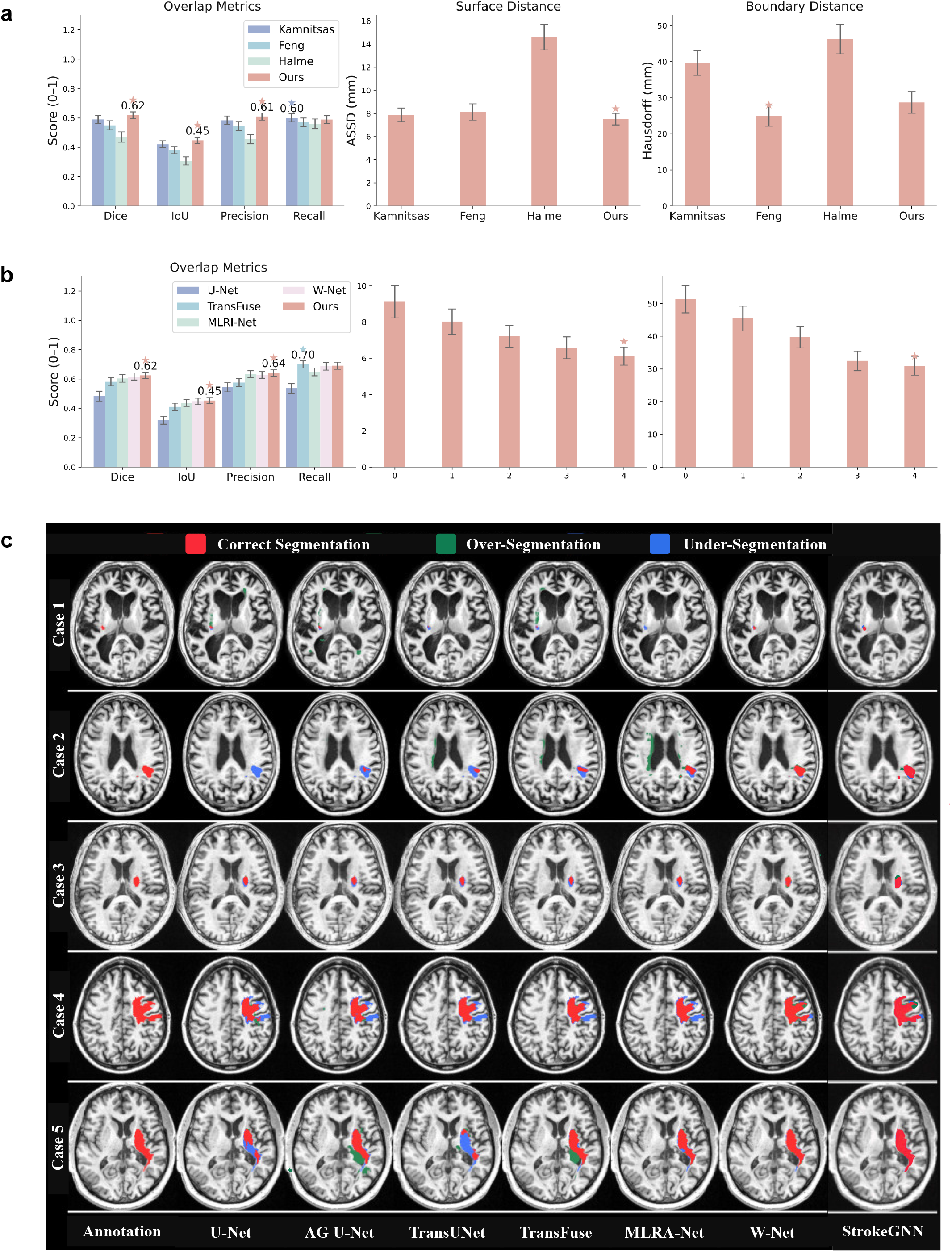
Experimental results for stroke lesion segmentation on MRI data. (a) Quantitative Comparison with existing state-of-the-art segmentation methods on SISS^35^. (b) Quantitative comparison with existing state-of-the-art segmentation methods on ATLAS v2.0. (c) Qualitative comparison with baseline models (U-net, TransFuse, MLRI-net, and W-net) on ATLAS v2.0 dataset.

We also compare our proposed method, StrokeGNN, with existing approaches on the ATLAS R2.0 dataset, including U-Net^21^, TransFuse^38^, MLRI-Net^39^, and W-Net^26^. Figure 3b (see Supplementary Figure S3d for details) presents the test results in terms of Dice score, precision, recall, and Hausdorff Distance (HD). As shown in Figure 3b, StrokeGNN outperforms the compared methods in Dice score, precision, and HD. In addition to the quantitative results, Figure 3c provides a qualitative comparison across five representative test cases. Our method consistently produces high-quality segmentation results. For Cases 1–3, which involve small lesion regions, StrokeGNN demonstrates substantial improvements by effectively capturing contextual information from adjacent slices. For Cases 4 and 5, it delivers accurate and well-defined segmentations, demonstrating both precision and robustness.

### Stroke lesion volume estimation from segmentation

To quantitatively assess lesion burden, we estimate ischemic stroke lesion volume using the same un-augmented MRI scans employed in the segmentation experiments (Figure 2a). Lesion volume is calculated directly from the predicted segmentation mask by multiplying the number of lesion voxels by the physical volume represented by each voxel, as described in Section . The estimated volume is then compared with the corresponding volume calculated from the ground-truth lesion mask.

Quantitatively, our framework achieves a relative error of 8.4% in lesion volume estimation, representing a ∼1.9% absolute reduction compared with baseline segmentation methods (Figure 4a-b). These results demonstrate that the proposed system not only delineates lesions accurately but also preserves volumetric fidelity, demonstrating that improved lesion delineation also translates into more accurate quantitative lesion-volume measurements.

**Figure 4.**
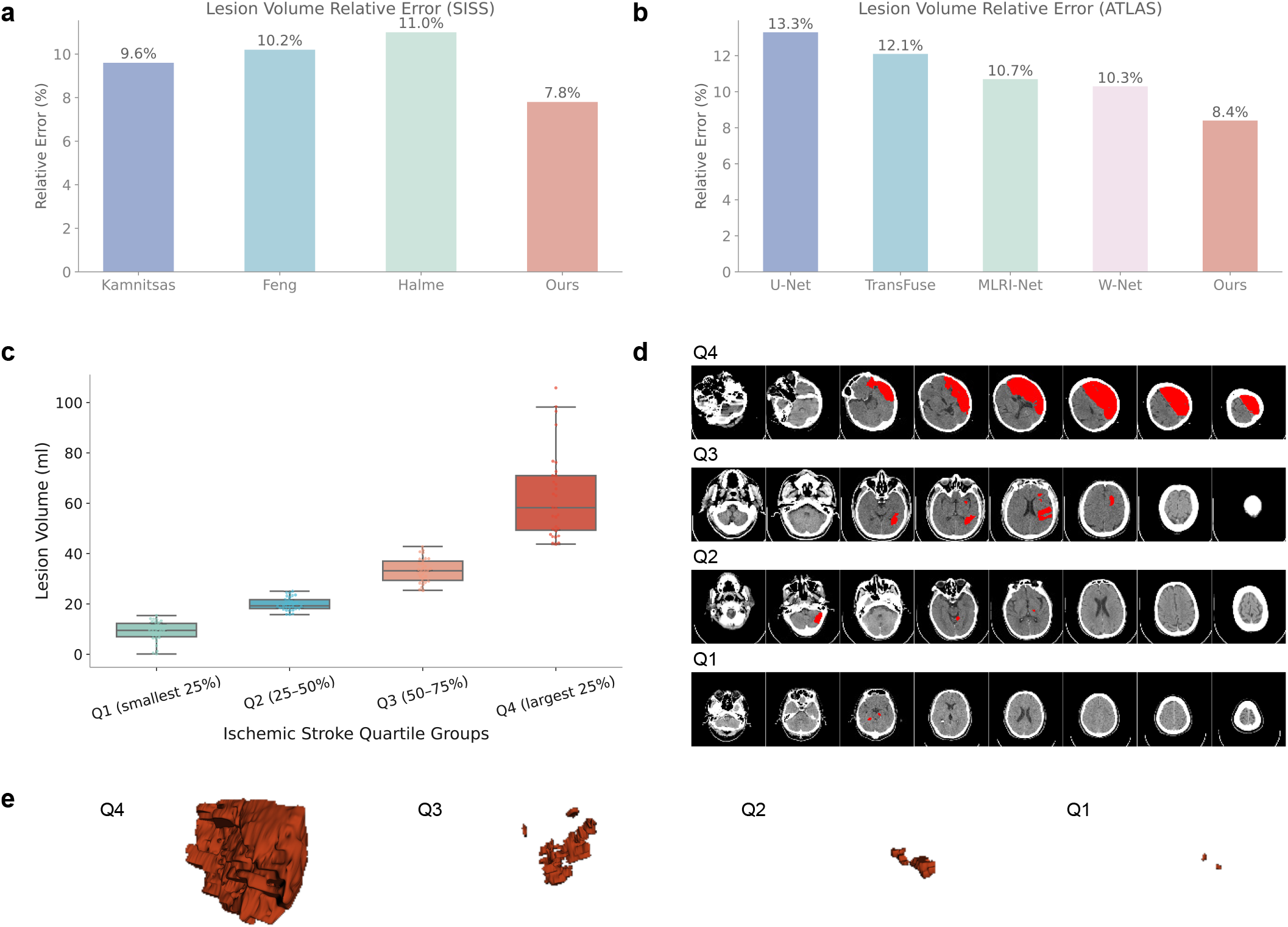
Experimental results for stroke lesion volume estimation. (a) Lesion volume relative error for SISS dataset. (b) Lesion volume relative error for ATLAS R2.0 Dataset. (c) Lesion volume groups. (c) Box plot of each cluster in terms of the segmented lesion volumes. (d) Sample MRI scans. (e) corresponding 3D visualization of Q1-Q4 lesion volume groups (segmented lesion is highlighted in red color).

**Figure 5.**
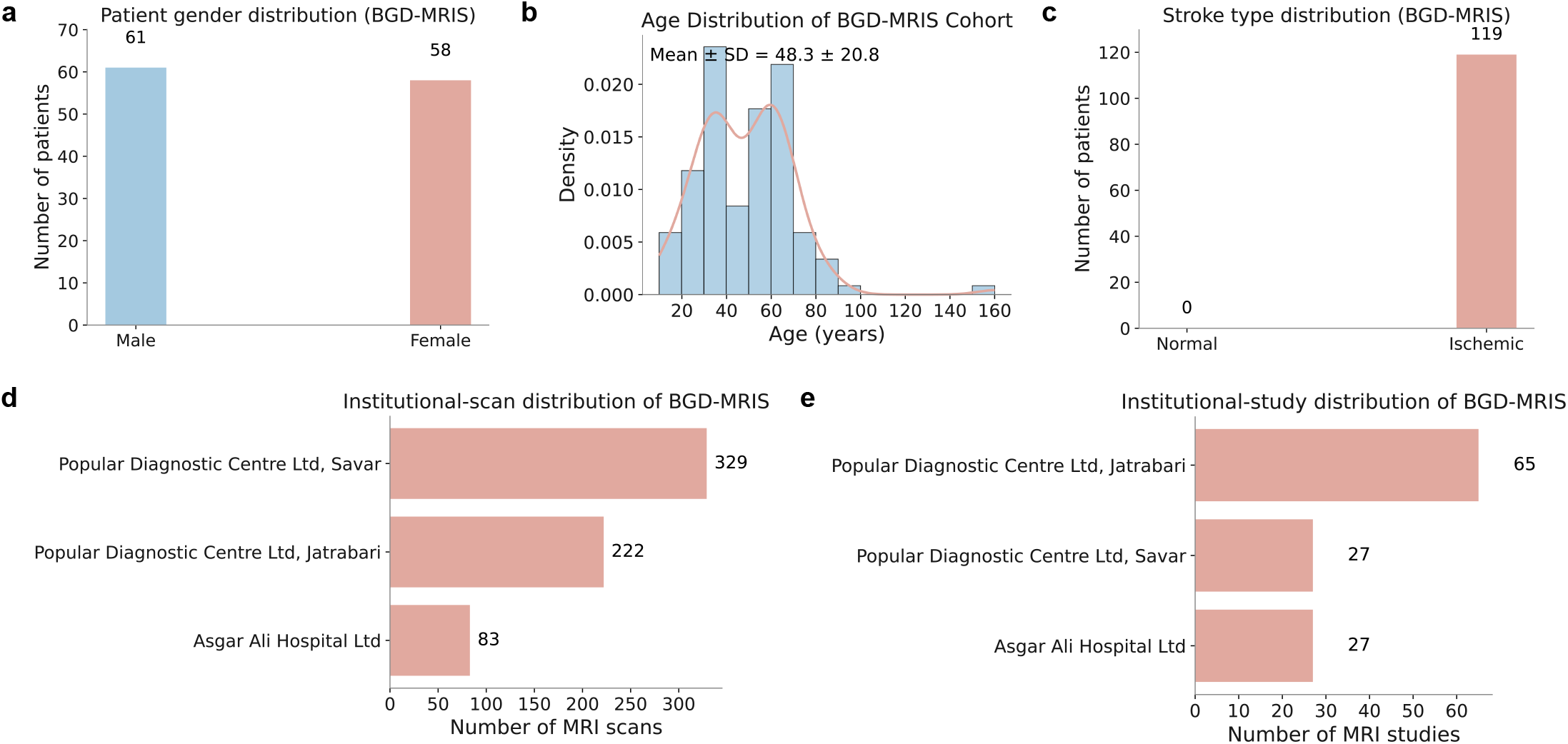
Demographic overview of the BGD-MRIS cohort. (a) Gender distribution of the enrolled subjects. (b) Age distribution of patients, shown as a histogram with density estimation (mean± s.d. indicated). (c) Distribution of ischemic stroke and control cases across the cohort. (d) Institutional distribution by the number of studies contributed from each participating medical centre. (e) Institutional distribution by number of MRI scans, reflecting inter-site variability in imaging volume.

For visualization and descriptive analysis, estimated lesion volumes are further divided into four groups using quartile thresholds (Q1–Q4), producing approximately equal-sized groups spanning the observed distribution of lesion burden. These quartiles are used only to illustrate increasing lesion volume and are not intended to represent clinical severity categories.

Figure 4c–g illustrate both quantitative and qualitative findings. Figure 4c presents the distribution of estimated lesion volumes across quartile groups for ischemic stroke populations, demonstrating a clear monotonic progression from Q1 to Q4. For qualitative validation, Figure 4d-e show representative axial MRI slices and corresponding 3D volumetric reconstructions from each quartile. These examples highlight the transition from small, well-circumscribed lesions in Q1 to extensive, multi-regional or diffuse abnormalities in Q4, underscoring the ability of MRI to capture fine-grained structural details.

Overall, this quartile-based visualization provides a simple and interpretable representation of increasing lesion burden from MRI. Combined with the low relative error in lesion volume estimation, these results demonstrate the feasibility of deriving quantitative lesion burden directly from automated segmentation. Future work will investigate the relationship between imaging-derived lesion characteristics, including volume, location, shape, and spatial distribution, and independently acquired clinical outcome measures.

### Extended evaluation of ischemic stroke detection

To develop and rigorously evaluate our ischemic stroke detection framework, we constructed the BGD-MRIS dataset, a multi-center brain MRI cohort collected from three tertiary hospitals in Bangladesh: Asgar Ali Hospital Ltd; Popular Diagnostic Centre Ltd (Jatrabari Branch, Dhaka); and Popular Diagnostic Centre Ltd (Savar). MRI scans were acquired between July 10, 2023 and June 14, 2024 under routine clinical protocols.

The dataset contains 532 MRI scans (21 ADC, 26 DWI, 5 FLAIR, 144 T1, and 336 T2) of 115 patients diagnosed with acute ischemic stroke. All cases were independently reviewed and annotated by two board-certified radiologists with expertise in neuroimaging. In cases of disagreement, consensus was reached through joint review. To further assess cross-institutional robustness and protocol variability, we additionally evaluate our model on the publicly available ISLES dataset, which contains multi-sequence MRI scans from international centers.

Unlike traditional classification networks trained directly from raw images, our detection module leverages segmentation-informed representations. Specifically, volumetric feature maps extracted from the trained lesion segmentation backbone are used as input to the detection head. Intermediate feature tensors of size R^32×32×32×256^ are globally pooled to produce a compact representation that preserves spatially aggregated lesion information. The final output layer produces a scalar probability corresponding to ischemic stroke presence (yes/no) via sigmoid activation. This design enables the detection network to inherit anatomically meaningful lesion-aware features, improving sensitivity to subtle ischemic patterns.

Model optimization is performed using binary cross-entropy loss. We employ stochastic gradient descent (SGD) with an initial learning rate of 0.001, momentum of 0.9, and weight decay of 0.001. Each MRI volume is resized to ℝ^32×256×256×1^ prior to feature extraction. Training is conducted with a batch size of 4. Early stopping is applied if validation loss does not improve for 10 consecutive epochs, preventing overfitting.

To ensure a statistically reliable evaluation, we adopt five-fold cross-validation. In each fold, 80% of the data are used for training and 20% for testing. From the training subset, 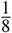is reserved for validation to guide model selection. Data augmentation, including spatial transformations and intensity perturbations, is applied exclusively to the training data to enhance generalization. All preprocessing and augmentation strategies are identical to those used in the segmentation experiments described in Section ISDS-MRI-enabled ischemic stroke lesion segmentation.

Across the five folds, model convergence is achieved after approximately 34 epochs on average (approximately 42 minutes per fold). Supplementary Figure S4a shows representative training and validation loss curves, demonstrating stable convergence without substantial divergence between the two curves. The optimal model is typically selected between epochs 24 and 30, with early stopping triggered around the 36^th^ epoch.

The overall detection performance is summarized in Figure 6. Figure 6a compares the proposed method with widely adopted 3D convolutional architectures, including 3D ResNet, 3D DenseNet, and 3D EfficientNet, across AUC, accuracy, precision, recall, specificity, and F1-score. All baseline models are trained under identical preprocessing, augmentation, and cross-validation settings to ensure a fair comparison. Our segmentation-guided detection framework achieves consistently strong performance across these metrics, with an average AUC of 0.962 for ischemic stroke detection. The corresponding ROC curves are presented in Figure 6b, further demonstrating the discriminative capability of the proposed method across different operating thresholds. Figure 6c further reports the performance of the proposed method across the five cross-validation folds. AUC, accuracy, recall, and specificity remain stable across folds, indicating that the observed performance is not driven by a particular data split. Specifically, specificity consistently exceeds 0.94, demonstrating reliable exclusion of lesion-negative samples, while recall remains high across all folds, indicating robust identification of lesion-positive samples. Detailed fold-wise results for AUC, accuracy, precision, recall, specificity, and F1-score are provided in Supplementary Figure S4b, while the corresponding fold-specific ROC curves are shown in Supplementary Figure S4c.

**Figure 6.**
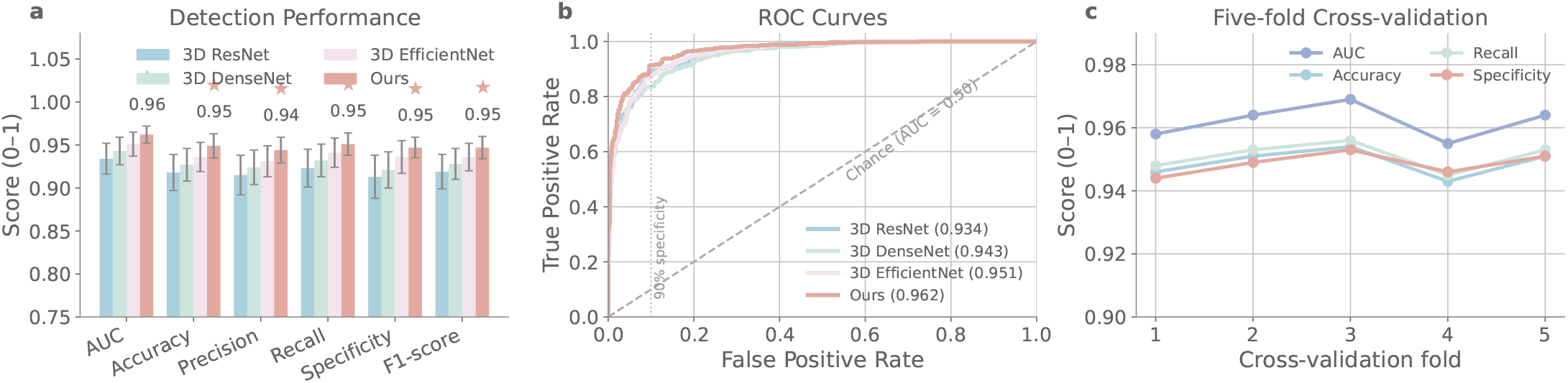
Evaluation of ischemic stroke detection. (a) Quantitative comparison of ISDS-MRI with 3D ResNet, 3D DenseNet and 3D EfficientNet across AUC, accuracy, precision, recall, specificity and F1-score. (b) Receiver operating characteristic curves of the comparison methods. (c) Detection performance across five cross-validation folds.

Collectively, these results demonstrate that incorporating segmentation-derived spatial representations into the ischemic stroke detection module provides a measurable advantage over conventional 3D classification architectures. The consistent quantitative performance, ROC analysis, and limited variation across cross-validation folds further support the robustness of the proposed approach for ischemic lesion detection from heterogeneous MRI data.

### Ablation Analysis of StrokeGNN for MRI Lesion Segmentation and Detection

We performed comprehensive ablation experiments to systematically assess the architectural and training choices underlying the proposed MRI-based StrokeGNN framework. The ablation experiments focus on the principal components underlying ischemic lesion segmentation, volume estimation, and lesion detection, including volumetric reasoning, graph construction, and feature transfer between segmentation and detection modules.

#### Segmentation Backbone and Volumetric Modelling

We first compared the proposed 2D encoder augmented with StrokeGNN layers against (i) a standard 2D U-Net and (ii) a conventional 3D U-Net. As reported in Supplementary Table S3a, incorporating StrokeGNN consistently improves Dice, IoU, precision, recall, and surface-distance metrics across datasets. Compared with the pure 2D backbone, the gains confirm the importance of modelling inter-slice dependencies. Relative to 3D convolutions, StrokeGNN achieves competitive or superior performance while avoiding excessive memory consumption and boundary artefacts, demonstrating the effectiveness of graph-based volumetric reasoning.

#### Effect of Graph Neighbourhood Size

We next investigated the influence of the graph neighbourhood parameter *k*. Results in Supplementary Table S3b show that performance improves when increasing *k* from 1 to 3, indicating that limited cross-slice aggregation enhances contextual understanding. However, larger neighbourhood sizes lead to marginal degradation, likely due to feature oversmoothing and reduced discriminability. These findings suggest that moderate local connectivity is sufficient to capture relevant volumetric context in ischemic lesion segmentation.

#### Number of StrokeGNN Layers

To examine the depth of graph reasoning, we evaluated configurations with one, two, and three StrokeGNN layers (Supplementary Table S3c). Two layers provide the best trade-off between contextual aggregation and optimization stability. Additional layers yield diminishing returns, suggesting that excessive message passing may not further benefit lesion delineation.

#### Patch-Level Lesion Detection

Since the MRI datasets used in this study contain only stroke-positive subjects, detection is evaluated at the patch level rather than the subject level. We compared (i) a standalone CNN trained directly for patch classification and (ii) a detection head built upon features transferred from the segmentation backbone. As shown in Supplementary Table S4a, the transfer-learning strategy consistently improves AUC, accuracy, and F1-score. This indicates that features optimized for lesion localization provide stronger discriminative cues for identifying lesion-containing patches than training a classifier from scratch.

#### Impact on Volume Estimation

Finally, we assessed how segmentation accuracy affects downstream lesion volume estimation. Supplementary Table S4b demonstrates that improvements in Dice score translate directly into reduced relative volume error. This confirms that precise boundary delineation is critical for reliable volumetric quantification.

Overall, these ablation experiments validate the principal design choices of the proposed MRI StrokeGNN framework, including graph-based volumetric reasoning, optimized neighbourhood size, appropriate graph depth, and segmentation-informed detection. The results collectively demonstrate that structured inter-slice modelling enhances both lesion delineation and downstream quantitative analysis.

## Discussion

AI-driven stroke diagnosis using MRI has substantial potential to improve clinical decision-making by enabling efficient and accurate lesion detection, segmentation, and quantitative lesion volume estimation within a unified computational framework. In this study, we present ISDS-MRI, a comprehensive MRI-based stroke diagnostic system that integrates lesion segmentation, and lesion volume estimation into a single end-to-end architecture. The framework further accommodates heterogeneous combinations of MRI sequences, allowing cases with incomplete sequence availability to be processed without requiring synthesis of missing images. The proposed framework demonstrates strong and consistent performance across multiple public datasets and our newly collected multi-center BGD-MRIS cohort, highlighting its robustness and generalizability.

**Lesion segmentation** serves as the foundation of MRI-based stroke analysis. Compared with CT, MRI provides richer soft-tissue resolution and improved sensitivity for early ischemic changes. Most existing approaches rely on conventional 3D convolutional neural networks that insufficiently model long-range anatomical dependencies. Our proposed StrokeGNN segmentation module enhances a U-Net backbone with a 3D graph neural network (GNN) to explicitly capture inter-slice and volumetric contextual relationships. This design enables the model to jointly exploit local intensity patterns and global structural continuity. Experimental results demonstrate that StrokeGNN consistently achieves superior Dice scores and precision compared with state-of-the-art 3D CNN baselines, while maintaining competitive recall. Qualitative analyses further confirm improved boundary delineation and robustness in cases involving small, diffuse, or multi-focal lesions.

**Stroke type detection** is clinically essential because ischemic and hemorrhagic strokes require fundamentally different treatment strategies. Thrombolytic therapy, for example, is indicated for ischemic stroke but contraindicated in hemorrhagic cases. While many prior studies focus on binary classification, our framework supports multi-class discrimination among ischemic stroke, hemorrhagic stroke, and non-stroke categories. By transferring volumetric feature representations learned during segmentation, the classification module leverages lesion-aware embeddings without redundant retraining. This feature-sharing strategy improves parameter efficiency and enhances generalization. Across cross-validation and external datasets, the model achieves high AUC values for both ischemic and hemorrhagic stroke detection, demonstrating stable performance across institutions and acquisition settings. Such robustness suggests potential utility in emergency settings where rapid MRI-based triage may be required.

**Lesion volume estimation** provides an additional layer of clinical relevance by estimating lesion burden and supporting treatment prioritization. Due to the scarcity of MRI datasets annotated with standardized severity scores, supervised severity prediction remains challenging. To address this limitation, we propose a quartile-based stratification framework that uses estimated lesion volumes as a quantitative proxy for severity. Patients are grouped into four interpretable categories spanning the full lesion distribution. Quantitative evaluation demonstrates low relative error in lesion volume estimation, supporting the reliability of downstream severity grouping. Visual analyses reveal clear monotonic progression in lesion extent and spatial involvement across quartiles. Although preliminary, this approach establishes a practical, label-free pathway for integrating unsupervised severity estimation into MRI-based stroke workflows.

From a systems perspective, ISDS-MRI is designed to be modular and clinically adaptable. Each component (segmentation, ischemic stroke detection, and lesion volume estimation) can function independently or as part of a unified pipeline. Segmentation outputs may be overlaid on radiological viewers for enhanced interpretability; detection outputs can indicate the presence of lesion-containing regions, while automated volumetric measurements can provide an objective quantitative description of lesion burden. Importantly, the framework operates under heterogeneous multi-site MRI conditions, demonstrating resilience to protocol variability and incomplete modality scenarios.

Nevertheless, several limitations warrant discussion. First, although the BGD-MRIS cohort improves demographic and institutional diversity, broader international validation will be necessary to confirm large-scale generalizability. Second, while quantitative metrics and visual analyses indicate strong performance, prospective clinical evaluation will ultimately be required to determine real-world impact. Failure cases suggest that extremely small lesions, severe motion artifacts, or pronounced intensity inhomogeneity may still challenge the model.

Future work will focus on expanding multi-center cohorts with consistently acquired MRI sequences and matched control subjects, further evaluating robustness to missing sequences, and prospectively validating the framework in clinical workflows. In summary, ISDS-MRI provides a scalable, interpretable, and clinically aligned framework for comprehensive MRI-based stroke diagnosis. Its unified design, strong empirical performance, and adaptability to heterogeneous imaging environments represent a meaningful step toward practical AI-assisted stroke care.

## Methods

To enable accurate ischemic stroke analysis using MRI, we propose ISDS-MRI, a unified framework that integrates lesion segmentation, ischemic stroke detection, and lesion volume estimation. Unlike CT-based systems that rely on Hounsfield unit normalization, MRI data exhibit modality-dependent intensity characteristics. Therefore, our pipeline is specifically designed to operate on multi-sequence MRI with standardized preprocessing. The overall architecture consists of a 2D encoder–decoder backbone augmented with StrokeGNN layers for volumetric reasoning, followed by task-specific heads for segmentation and detection. Detailed implementation settings are summarized in Supplementary Figure S1. To accommodate missing MRI sequences, each sequence is processed by an independently trained sequence-specific branch. For a given case, only branches corresponding to the available sequences are activated, and their outputs are aggregated to obtain the final prediction. Therefore, the framework does not require a fixed sequence combination across patients.

### StrokeGNN layer

Conventional 3D convolutional layers model volumetric context using fixed receptive fields. However, their performance may degrade at boundary slices when padding is applied to preserve spatial resolution. To address this limitation, we introduce StrokeGNN, a graph-based module that explicitly models inter-slice dependencies.

Given an intermediate MRI feature tensor of size 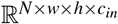, where *N* denotes the number of slices and *w*× *h* the in-plane spatial dimensions, each slice is treated as a graph node:

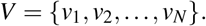

For each node *v*_*i*_, we define its neighbourhood using *k*-nearest neighbours in feature space:

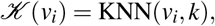

with *k* = 3 in this work. The adjacency matrix *A* ∈ {0, 1}^*N*×*N*^ is defined as

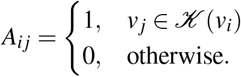

Feature propagation follows a message-passing mechanism. Neighbour messages are aggregated as

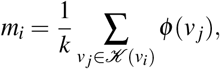

where *φ* (·) denotes a learnable 3 × 3 convolution. The updated node representation is

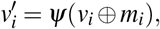

where *ψ*(·) is another learnable convolution and ⊕ denotes concatenation.

ReLU activation is applied after each convolution:

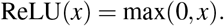

This design enables adaptive cross-slice information exchange while preserving in-plane spatial resolution.

### Stroke Lesion Segmentation

All MRI scans are resampled to a uniform in-plane resolution of 0.5× 0.5 mm while preserving the original through-plane spacing:

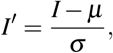

where *µ* and *σ* denote the mean and standard deviation of each volume.

The input MRI volume has size ℝ^*D*×*W* ×*H*×1^. To reduce memory usage, volumes are divided into patches of size ℝ^*d*×*w*×*h*×1^. Each patch is processed by a 2D U-Net encoder–decoder backbone with skip connections. The encoder extracts in-plane features, producing a tensor of size ℝ^*d*×*w*×*h*×64^.

Two StrokeGNN layers are then applied along the slice dimension to incorporate volumetric context. A final 1×1 convolution followed by sigmoid activation produces a binary lesion probability map:

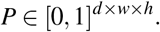

During inference, overlapping patch predictions are merged by averaging probabilities:

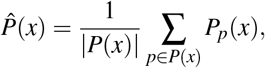

where *P*(*x*) denotes the set of patches covering voxel *x*.

The segmentation loss combines Dice and binary cross-entropy terms:

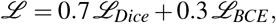

with

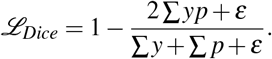

The model is optimized using RMSprop with learning rate 1×10^−5^ and early stopping based on validation Dice score.

### Stroke Lesion Volume Estimation

Following segmentation, lesion volume is computed directly from the predicted 3D binary mask. Let *M*∈ {0, 1} ^*D*×*W*×*H*^ denote the final lesion mask and (*s*_*d*_, *s*_*w*_, *s*_*h*_) the voxel spacing obtained from MRI metadata. The lesion volume is calculated as

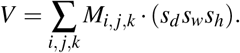

Equivalently, if *N*_*vox*_ denotes the number of lesion voxels,

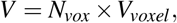

where *V*_*voxel*_ = *s*_*d*_*s*_*w*_*s*_*h*_.

This direct voxel-counting approach avoids geometric approximations and ensures that volume estimation remains fully consistent with the segmentation output. No clustering or severity categorisation is performed in this MRI study; volume serves solely as a quantitative biomarker derived from the predicted mask.

The relative volume error is calculated as

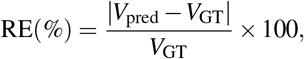

where *V*_pred_ and *V*_GT_ denote lesion volumes calculated from the predicted and ground-truth masks, respectively.

### Ischemic Stroke Detection

Unlike the CT framework that performs multi-class stroke classification, this MRI study focuses exclusively on binary ischemic stroke detection (yes/no).

We leverage transfer learning by reusing high-level features from the segmentation backbone. Let 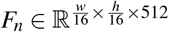 denote slice-level bottleneck features. Global average pooling produces:

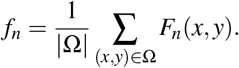

To aggregate slice-level features into a patch-level representation, attention pooling is applied:

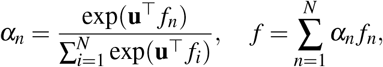

where **u** is a learnable attention vector.

The final feature vector *f* ∈ℝ^512^ is passed through two fully connected layers with dropout (0.1), followed by a sigmoid activation producing probability *p*∈ [0, 1] indicating ischemic stroke presence.

Binary cross-entropy loss is used:

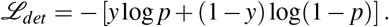

Training is performed using SGD with learning rate 1×10^−3^, momentum 0.9, weight decay 0.001, batch size 2, and early stopping if validation loss does not improve for 10 consecutive epochs.

### Implementation details

The training environment consists of a workstation powered by an Intel Core i7-13700 processor and an NVIDIA RTX 4090 GPU. It includes 32GB of RAM and operates on Ubuntu 20.04. The proposed models are developed in PyTorch, utilizing CUDA

11.8 to leverage GPU acceleration.

## Data Availability

All data produced will be available online at https://github.com/Zhicheng-Lu/stroke_mri upon the acceptance of the paper.

## Data availability

The public datasets used in this study are available from their respective repositories. The BGD-MRIS dataset will be made publicly available at:

BGD-MRIS: https://github.com/Zhicheng-Lu/stroke_mri.

SISS & SPES: https://www.smir.ch/ISLES/Start2015.

ISLES22: https://isles22.grand-challenge.org/.

ATLAS R2.0: https://atlas.grand-challenge.org/Data/.

## Code availability

All source codes are publicly available at https://github.com/Zhicheng-Lu/stroke_mri.

## Acknowledgments

This work was supported by Australian Commonwealth Funding.

## Author contributions statement

Z.Lu, S.Uddin, S.Uribe, S.White, R.T.Martins, S.Chau, A.S.M.Mosaddek, M.S.Islam, N.Nahar, A.K.M.Azad, K.M.N.Hossain, H.S.Choudhury, K.M.R.Hasan, S.Aloteibi, N.Mosaddek, S.Rahman, M.M.Hossain, and K.M.M.H.Sizar, P.Liò, M.T.Islam, and M.A.Moni made contributions to the concept and design of the article. Z.Lu conceived the study, designed the methodology, developed the models, conducted the experiments, and analysed the results. A.S.M.Mosaddek, M.S.Islam, N.Nahar, K.M.N.Hossain, H.S.Choudhury, K.M.R.Hasan, N.Mosaddek, S.Rahman, M.M.Hossain, and K.M.M.H.Sizar contributed to data curation and clinical interpretation. S.Uribe, S.White, R.T.Martins, A.K.M.Azad, S.Chau, C.Angione provided domain expertise and guidance on imaging protocols. M.A.Moni supervised the project. All authors reviewed and approved the manuscript.

