## Supplementary for "Ischemic Stroke Detection, Segmentation, and Volume Estimation using Multi-sequence MRI Data with Missing Sequences"

#### This PDF file includes:

Figs. S1 to S4
Tables S1 to S6
SI References

### Demographic Information of Curated BGD-MRIS

**Table S1. Demography overview of the collected BGD-MRIS. (a) Patient-level characteristics, including age distribution, gender ratio, and stroke type breakdown. (b) Institutional distribution of the dataset across contributing medical centres.**

| Characteristic | Value |
|---|---|
| Total subjects | 119 |
| Age (mean $\pm$ SD) | 48.3 $\pm$ 20.8 years |
| Age range | 10 - 90 |
| Gender (M / F) | 61 (51.3%) / 58 (48.7%) |
| Stroke type | 119 ischemic (100%) |

**(a)**

| Institution | Studies | Scans |
|---|---|---|
| Asgar Ali Hospital Ltd | 27 | 83 |
| Popular Diagnostic Centre Ltd, Jatrabari Branch Dhaka | 65 | 222 |
| Popular Diagnostic Centre Ltd, Savar | 27 | 329 |
| **Total** | 597 | 1,507 |

**(b)**

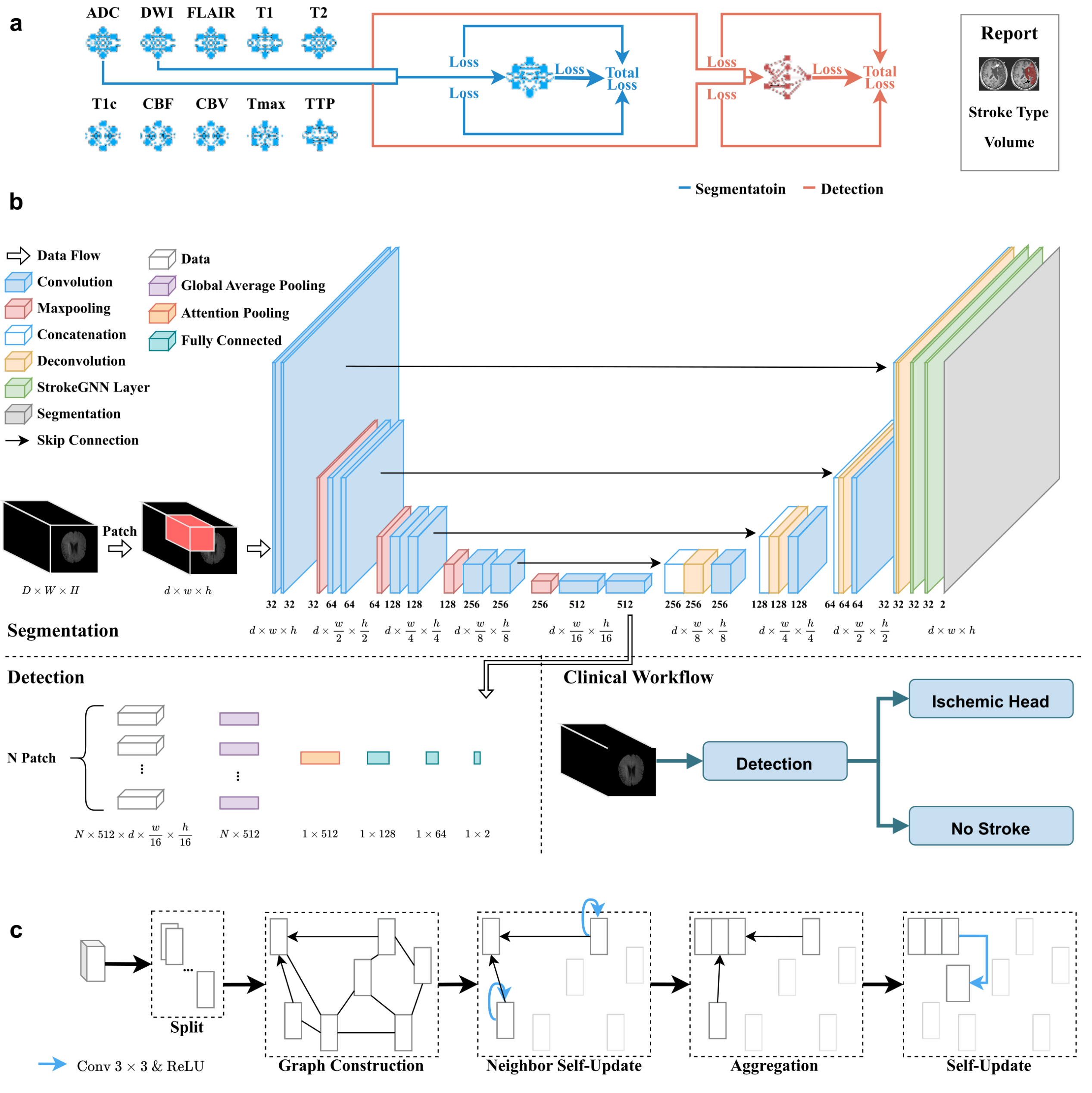


(a)

**Fig. S1.** Details of the proposed ISDS-MRI including sub-models and GNN layer. (a) Overview of multiple sub-models design for multi-modal MRI data, we use ISLES22 dataset with ADC and DWI sequences as example. For each of the segmentation (blue) and detection (orange), total loss is calculated from the average of individual sub-models results and integrated results. Details of each sub-models are shown in (b). (b) Architecture of ISDS-MRIfor ischemic stroke analysis from multi-modal MRI. Each MRI sequence is processed by a modality-specific sub-network based on a U-Net architecture, producing voxel-wise ischemic lesion segmentation. Bottleneck features learned during segmentation are further reused for downstream tasks, including stroke detection and lesion volume estimation. Predictions from available modalities are aggregated to generate final case-level and voxel-level outputs, enabling robust operation under missing-modality conditions. (c) Illustration of the StrokeGNN layer. Three-dimensional MRI volumes are decomposed into two-dimensional slices, with each slice represented as a node in a graph. Edges are constructed by connecting each node to its $K$ nearest neighbors in the feature space. Graph convolution enables each node to aggregate contextual information from semantically similar slices, enhancing the modeling of three-dimensional spatial dependencies.

**Table S2. Proposed model details for each MRI sequence. (a) Stroke segmentation module details. All convolutional layers use $1 \times 1$ stride and "same" padding, and apply ReLU activation function afterwards except final "Conv" layer. (b) Stroke detection module details. Input is from Table S2a Up1_2 layer. All convolutional and fully connected layers use $1 \times 1$ stride and "same" padding, and apply ReLU activation function afterwards except final "FC3" layer.**

| | | |
|---|---|---|
| Down1 | Down1_1 | Input: $\mathbb{R}^{1\times d\times w\times h}$. Kernel: $3 \times 3$. Out_channel : 32. |
| | Down1_2 | Kernel: $3 \times 3$. Out_channel: 32. |
| | Maxpool1 | Stride: $2 \times 2$. |
| Down2 | Down2_1 | Input: Maxpool1 $\in \mathbb{R}^{32\times d\times \frac{w}{2}\times \frac{h}{2}}$. Kernel: $3 \times 3$. Out_channel: 64. |
| | Down2_2 | Kernel: $3 \times 3$. Out_channel: 64. |
| | Maxpool2 | Stride: $2 \times 2$. |
| Down3 | Down3_1 | Input: Maxpool2 $\in \mathbb{R}^{64\times d\times \frac{w}{4}\times \frac{h}{4}}$. Kernel: $3 \times 3$. Out_channel: 128. |
| | Down3_2 | Kernel: $3 \times 3$. Out_channel: 128. |
| | Maxpool3 | Stride: $2 \times 2$. |
| Down4 | Down4_1 | Input: Maxpool3 $\in \mathbb{R}^{128\times d\times \frac{w}{8}\times \frac{h}{8}}$. Kernel: $3 \times 3$. Out_channel: 256. |
| | Down4_2 | Kernel: $3 \times 3$. Out_channel: 256. |
| | Maxpool4 | Stride: $2 \times 2$. |
| Up1 | Up1_1 | Input: Maxpool4 $\in \mathbb{R}^{256\times d\times \frac{w}{16}\times \frac{h}{16}}$. Kernel: $3 \times 3$. Out_channel: 512. |
| | Up1_2 | Kernel: $3 \times 3$. Out_channel: 512. |
| | UpConv1 | Kernel: $3 \times 3$. Stride: $2 \times 2$. Out_channel:256. |
| Up2 | Concat1 | Input: Down4_2 $\in \mathbb{R}^{256\times d\times \frac{w}{8}\times \frac{h}{8}}$, UpConv1 $\in \mathbb{R}^{256\times d\times \frac{w}{8}\times \frac{h}{8}}$. Axis: 0. |
| | Up2_1 | Kernel: $3 \times 3$. Out_channel: 256. |
| | Up2_2 | Kernel: $3 \times 3$. Out_channel: 256. |
| | UpConv2 | Kernel: $3 \times 3$. Stride: $2 \times 2$. Out_channel: 128. |
| | Concat2 | Input: Down3_2 $\in \mathbb{R}^{128\times d\times \frac{w}{4}\times \frac{h}{4}}$, UpConv2 $\in \mathbb{R}^{128\times d\times \frac{w}{4}\times \frac{h}{4}}$. Axis: 0. |

| | | |
|---|---|---|
| Up3 | Up3_1 | Kernel: $3 \times 3$. Out_channel: 128. |
| | Up3_2 | Kernel: $3 \times 3$. Out_channel: 128. |
| | UpConv3 | Kernel: $3 \times 3$. Stride: $2 \times 2$. Out_channel: 64. |
| Up4 | Concat3 | Input: Down2_2 $\in \mathbb{R}^{64\times d\times \frac{w}{2}\times \frac{h}{2}}$, UpConv3 $\in \mathbb{R}^{64\times d\times \frac{w}{2}\times \frac{h}{2}}$. Axis: 0. |
| | Up4_1 | Kernel: $3 \times 3$. Out_channel: 64. |
| | Up4_2 | Kernel: $3 \times 3$. Out_channel: 64. |
| | UpConv4 | Kernel: $3 \times 3$. Stride: $2 \times 2$. Out_channel: 32. |
| | Concat4 | Input: Down1_2 $\in \mathbb{R}^{32\times d\times w\times h}$, UpConv4 $\in \mathbb{R}^{32\times d\times w\times h}$. Axis: 0. |
| GNN1_ KNN | Build1 | Input: Concat4 $\in \mathbb{R}^{64\times d\times w\times h}$. Output: $\mathbb{R}^{3\times 64\times d\times w\times h}$. |
| | Aggr1 | Kernal: $3 \times 3$. Out_channel: 64. |
| | Mean1 | Axis: 0. |
| GNN1 | Concat5 | Input: Concat4 $\in \mathbb{R}^{64\times d\times w\times h}$, Mean1 $\in \mathbb{R}^{64\times d\times w\times h}$. Axis: 0. |
| | Update1 | Kernel: $3 \times 3$. Out_channel: 64. |
| GNN2_ KNN | Build2 | Input: Update1 $\in \mathbb{R}^{64\times d\times w\times h}$. Output: $\mathbb{R}^{3\times 64\times d\times w\times h}$. |
| | Aggr2 | Kernal: $3 \times 3$. Out_channel: 64. |
| | Mean2 | Axis: 0. |
| GNN2 | Concat6 | Input: Update1 $\in \mathbb{R}^{64\times d\times w\times h}$, Mean2 $\in \mathbb{R}^{64\times d\times w\times h}$. Axis: 0. |
| | Update2 | Kernel: $3 \times 3$. Out_channel: 64. |
| | Conv | Kernel: $1 \times 1$. Out_channel: 2. |
| | Softmax | Axis: 0. |

**(a)**

| | | |
|---|---|---|
| Average | Patches | Input: Up1_2 $\in \mathbb{R}^{512\times d\times \frac{w}{16}\times \frac{h}{16}}$. Output: $\mathbb{R}^{N\times 512\times d\times \frac{w}{16}\times \frac{h}{16}}$. |
| | G_Average | Out_features: $N \times 512$. |
| Attention | A_Pooling | Input: G_Average $\in \mathbb{R}^{N\times 512}$. Output: $\mathbb{R}^{1\times 512}$ |
| FC1 | FC1 | Input: A_Pooling $\in \mathbb{R}^{1\times 512}$. Out_channel: 128. |
| | Dropout2 | p: 0.1. |
| FC2 | FC2 | Input: Dropout1 $\in \mathbb{R}^{1\times 128}$. Out_channel: 164. |
| | Dropout2 | p: 0.1. |
| FC3 | FC3 | Input: Dropout2 $\in \mathbb{R}^{1\times 64}$. Out_channel: 3. |
| | Softmax | Axis: 0. |

**(b)**

### Supplementary: Segmentation

| | ADC | DWI | FLAIR | T1 | T2 | T1c | CBF | CBV | Tmax | TTP |
|---|---|---|---|---|---|---|---|---|---|---|
| SISS (1) | - | 28 (840) | 28 (840) | 28 (840) | 28 (840) | - | - | - | - | - |
| SPES (1) | - | 30 (600) | - | - | 30 (600) | 30 (600) | 30 (600) | 30 (600) | 30 (600) | 30 (600) |
| ISLES22 (2) | 250 (750) | 250 (750) | - | - | - | - | - | - | - | - |
| ATLAS R2.0 (3) | - | - | - | 655 | - | - | - | - | - | - |
| **Total** | 250 (750) | 308 (2190) | 28 (840) | 683 (1495) | 58 (1440) | 30 (600) | 30 (600) | 30 (600) | 30 (600) | 30 (600) |

**(a)**

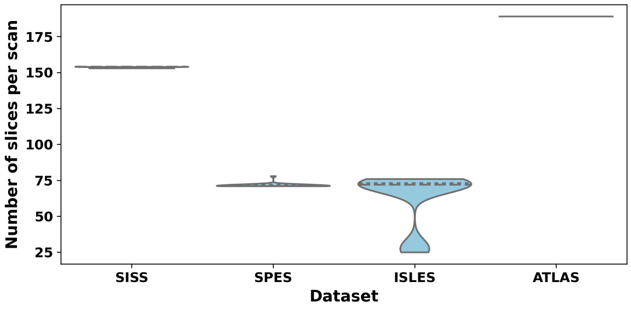


**(b)**

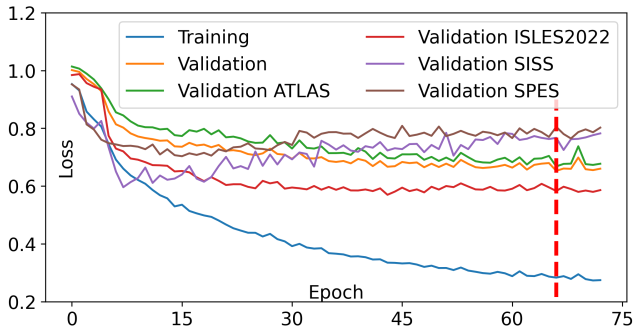


**(c)**

| | Dice ↑ | IoU ↑ | Precision ↑ | Recall ↑ | ASSD ↓ | Hausdorff ↓ |
|---|---|---|---|---|---|---|
| SISS | $60.77 \pm 2.8$ | $44.0 \pm 2.4$ | $58.9 \pm 3.1$ | $59.21 \pm 3.0$ | $7.33 \pm 0.6$ | $30.77 \pm 3.2$ |
| SPES | $83.88 \pm 1.9$ | $72.4 \pm 2.1$ | $82.1 \pm 2.3$ | $85.24 \pm 2.0$ | $\mathbf{2.34 \pm 0.3}$ | $\mathbf{10.73 \pm 2.1}$ |
| ISLES22 | $\mathbf{85.11 \pm 1.7}$ | $\mathbf{74.1 \pm 2.0}$ | $\mathbf{83.5 \pm 2.1}$ | $\mathbf{86.64 \pm 1.8}$ | $7.06 \pm 0.7$ | $28.48 \pm 3.4$ |
| ATLAS R2.0 | $62.69 \pm 2.6$ | $45.7 \pm 2.3$ | $61.0 \pm 2.8$ | $63.01 \pm 2.9$ | $7.65 \pm 0.8$ | $29.30 \pm 3.6$ |
| **Overall** | $72.50 \pm 2.2$ | $59.1 \pm 2.2$ | $71.4 \pm 2.4$ | $72.81 \pm 2.3$ | $6.28 \pm 0.5$ | $25.60 \pm 2.9$ |

**(d)** Cross-dataset segmentation performance of ISDS-MRI on public MRI datasets. Mean $\pm$ standard deviation is reported. Best performance per metric is shown in bold.

**Fig. S2.** Experimental results for stroke lesion segmentation in 5-fold cross validation. (a) Dataset distributions before and after augmentation. (b) Training and validation losses by epochs. (c) Quantitative testing results including Dice, IoU, precision, recall, ASSD, and Hausdorff distance across original datasets.

| | Training | Validation | Testing |
|---|---|---|---|
| SISS | 23 (690) | 5 (150) | 36 |
| SPES | 25 (500) | 5 (100) | 20 |
| ISLES22 | 200 (600) | 50 (150) | 150 |
| ATLAS R2.0 | 500 | 155 | 616 |
| **Total** | 748 (2290) | 215 (555) | 822 |

**(a)**

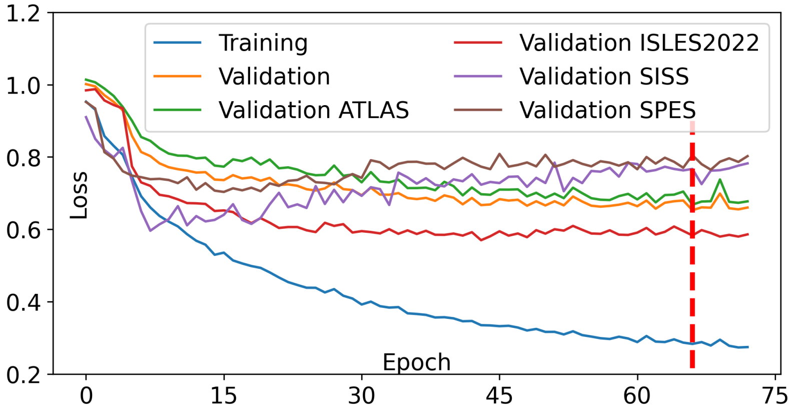


**(b)**

| | Dice (%) | IoU (%) | Precision (%) | Recall (%) | ASSD | HD |
|---|---|---|---|---|---|---|
| Kamnitsas *et al.* (4) | $59.0 \pm 2.7$ | $42.1 \pm 2.3$ | $58.4 \pm 2.9$ | $\mathbf{60.0 \pm 2.8}$ | $7.87 \pm 0.6$ | $39.61 \pm 3.4$ |
| Feng *et al.* (5) | $55.0 \pm 3.1$ | $38.1 \pm 2.5$ | $54.3 \pm 3.0$ | $57.0 \pm 3.0$ | $8.13 \pm 0.7$ | $\mathbf{25.02 \pm 2.9}$ |
| Halme *et al.* (6) | $47.0 \pm 3.5$ | $30.7 \pm 2.8$ | $45.6 \pm 3.2$ | $56.0 \pm 3.3$ | $14.61 \pm 1.1$ | $46.26 \pm 4.1$ |
| **Ours** | $\mathbf{61.82 \pm 2.3}$ | $\mathbf{44.7 \pm 2.1}$ | $\mathbf{60.9 \pm 2.4}$ | $58.94 \pm 2.6$ | $\mathbf{7.51 \pm 0.5}$ | $28.70 \pm 3.0$ |

**(c)**

| | Dice (%) | IoU (%) | Precision (%) | Recall (%) | ASSD | HD |
|---|---|---|---|---|---|---|
| U-Net (7) | $48.34 \pm 3.4$ | $31.9 \pm 2.7$ | $54.45 \pm 3.1$ | $53.68 \pm 3.2$ | $9.12 \pm 0.9$ | $51.35 \pm 4.2$ |
| TransFuse (8) | $58.18 \pm 2.9$ | $41.0 \pm 2.5$ | $57.64 \pm 2.7$ | $\mathbf{70.06 \pm 2.5}$ | $8.02 \pm 0.7$ | $45.44 \pm 3.8$ |
| MLRI-Net (9) | $60.48 \pm 2.6$ | $43.6 \pm 2.3$ | $63.29 \pm 2.4$ | $64.88 \pm 2.7$ | $7.21 \pm 0.6$ | $39.73 \pm 3.3$ |
| W-Net (10) | $61.76 \pm 2.4$ | $44.8 \pm 2.2$ | $62.86 \pm 2.3$ | $68.68 \pm 2.6$ | $6.58 \pm 0.6$ | $32.47 \pm 3.0$ |
| **Ours** | $\mathbf{62.44 \pm 2.1}$ | $\mathbf{45.4 \pm 2.0}$ | $\mathbf{64.17 \pm 2.2}$ | $69.03 \pm 2.4$ | $\mathbf{6.12 \pm 0.5}$ | $\mathbf{30.89 \pm 2.8}$ |

**(d)**

**Fig. S3.** Experimental results for stroke lesion segmentation on MRI data. (a) Dataset distribution for lesion segmentation testing. Note that numbers in the brackets are the dataset size after augmentation. (b) Training and validation loss values by epoch with setup described in Table S3a. (c) Quantitative Comparison with existing state-of-the-art segmentation methods on SISS. (d) Quantitative comparison with existing state-of-the-art segmentation methods on ATLAS R2.0. (e) Qualitative comparison with baseline models (U-net, TransFuse, MLRI-net, and W-net) on ATLAS R2.0 dataset.

### Supplementary: Ischemic Stroke Detection

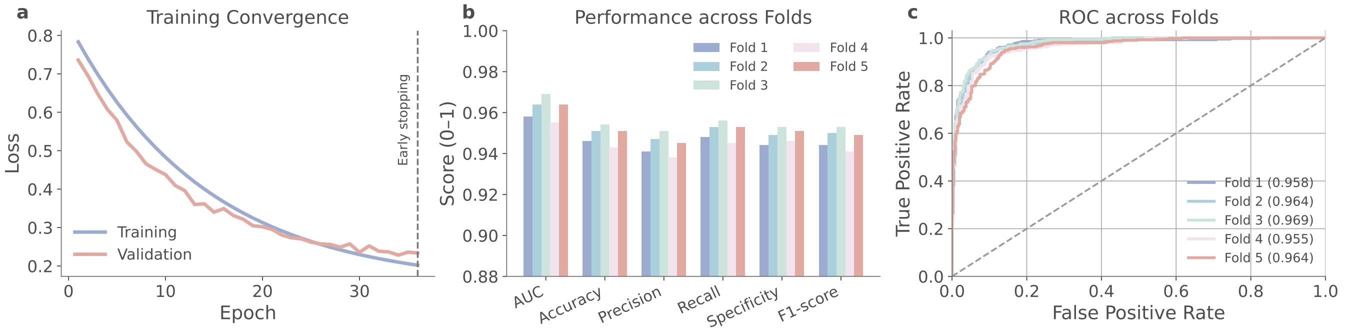


**Fig. S4.** Detailed evaluation of ischemic stroke detection. (a) Representative training and validation loss curves. (b) AUC, accuracy, precision, recall, specificity and F1-score across the five cross-validation folds. (c) Receiver operating characteristic curves for individual folds.

### Supplementary: Ablation Study

**Table S3. Ablation study for MRI segmentation module. (a) 2D U-Net vs 3D U-Net vs StrokeGNN. (b) Effect of graph neighbourhood size $k$. (c) Effect of number of StrokeGNN layers.**

| | Dice ↑ | IoU ↑ | Precision ↑ | Recall ↑ | ASSD ↓ | Hausdorff ↓ |
|---|---|---|---|---|---|---|
| 2D U-Net (7) | 0.7012 | 0.5480 | 0.7245 | 0.6891 | 1.9 | 7.6 |
| 3D U-Net (7) | 0.7148 | 0.5637 | 0.7410 | 0.7034 | 1.8 | 7.3 |
| **StrokeGNN** | **0.7426** | **0.5983** | **0.7689** | **0.7214** | **1.6** | **6.8** |

**(a)**

| | Dice ↑ | IoU ↑ | Precision ↑ | Recall ↑ | ASSD ↓ | Hausdorff ↓ |
|---|---|---|---|---|---|---|
| $k = 1$ | 0.7325 | 0.5841 | 0.7512 | 0.7180 | 1.7 | 7.1 |
| $k = 2$ | 0.7398 | 0.5936 | 0.7644 | 0.7193 | 1.6 | 6.9 |
| $\mathbf{k = 3}$ | **0.7426** | **0.5983** | **0.7689** | **0.7214** | **1.6** | **6.8** |
| $k = 4$ | 0.7359 | 0.5890 | 0.7602 | 0.7095 | 1.8 | 7.4 |
| $k = 5$ | 0.7210 | 0.5702 | 0.7425 | 0.6988 | 2.0 | 7.9 |

**(b)**

| | Dice ↑ | ASSD ↓ | Hausdorff ↓ |
|---|---|---|---|
| One StrokeGNN layer | 0.7364 | 1.7 | 7.0 |
| **Two StrokeGNN layers** | **0.7426** | **1.6** | **6.8** |
| Three StrokeGNN layers | 0.7391 | 1.7 | 7.2 |

**(c)**

**Table S4. Ablation study for MRI patch-level lesion detection and volume estimation. (a) Standalone CNN vs segmentation-feature transfer. (b) Impact of segmentation quality on relative volume error.**

| | AUC ↑ | Accuracy ↑ | Precision ↑ | Recall ↑ | F1-score ↑ |
|---|---|---|---|---|---|
| Standalone CNN | 0.9025 | 0.8764 | 0.8617 | 0.8842 | 0.8728 |
| **Segmentation feature transfer** | **0.9348** | **0.9013** | **0.8921** | **0.9105** | **0.9012** |

**(a)**

| | Dice ↑ | Relative Volume Error (%) ↓ |
|---|---|---|
| 2D U-Net | 0.7012 | 18.6 |
| 3D U-Net | 0.7148 | 16.9 |
| **StrokeGNN** | **0.7426** | **13.4** |

**(b)**

### Supplementary: Overview of Public Datasets

| Dataset Name | Download Link |
|---|---|
| SISS | https://www.smir.ch/ISLES/Start2015D |
| SPES | https://www.smir.ch/ISLES/Start2015 |
| ISLES22 | https://isles22.grand-challenge.org/ |
| ATLAS R2.0 | https://atlas.grand-challenge.org/Data/ |

**Table S5. Brief overview of public datasets used in this article. More detailed overviews are shown on the next page.**

| Acronym | Full Name | Download | Publication / Challenge | Size | Description |
|---|---|---|---|---|---|
| SISS | Stroke Imaging Segmentation Set | https://www.smir.ch/ISLES/Start2015D | IELES 2015 - A Public Evaluation Benchmark for Ischemic Stroke Lesion Segmentation from Multispectral MRI | 64 | The SISS dataset is part of the ISLES 2015 challenge and consists of multi-moda[illegible] MRI scans from patients with acute ischemic stroke. The dataset includes diffusion-weighted imaging (DWI), apparent diffusion coefficient (ADC), and perfusion-related sequences, along with expert-annotated ischemic lesion mask[illegible] It is commonly used for benchmarking ischemic stroke lesion segmentation algorithms. |
| SPES | Stroke Perfusion Estimation Set | https://www.smir.ch/ISLES/Start2015 | IELES 2015 - A Public Evaluation Benchmark for Ischemic Stroke Lesion Segmentation from Multispectral MRI | 50 | The SPES dataset, released as part of the ISLES 2015 challenge, focuses on ischemic stroke perfusion analysis. It contains multi-modal MRI data, including DWI, ADC, and perfusion maps, with lesion annotations derived from follow-up imaging. The dataset supports research on lesion outcome prediction and perfusion-informed stroke assessment. |
| ISLES22 | Ischemic Stroke Lesion Segmentation 2022 | https://isles22.grand-challenge.org/ | ISLES 2022: A Multi-Center Magnetic Resonance Imaging Stroke Lesion Segmentation Dataset | 400 | ISLES22 is a large-scale public dataset for ischemic stroke lesion segmentation from acute multi-modal MRI. It includes DWI and ADC sequences acquired withi[illegible] the hyperacute phase of stroke, with high-quality voxel-level annotations provide[illegible] by expert neuroradiologists. The dataset emphasizes robustness across institutions and scanner variability. |
| ATLAS R2.0 | Anatomical Tracings of Lesions After Stroke (v2.0) | https://atlas.grand-challenge.org/Data/ | A Large, Curated, Open-source Stroke Neuroimaging Dataset ot Improve Lesion Segmentation Algorithms | 1271 | ATLAS R2.0 is a large, heterogeneous MRI dataset containing manually segmented stroke lesions, primarily derived from T1-weighted images. The datase[illegible] includes chronic ischemic stroke cases collected across multiple sites and scanners, with annotations validated by trained experts. ATLAS is widely used fo[illegible] studying lesion anatomy, variability, and generalization in stroke segmentation models. |

**Table S6. Detailed overview of public MRI datasets used in this study. All datasets focus on ischemic stroke and provide expert-annotated lesion masks for segmentation and downstream analysis.**
